# Validation and performance evaluation of a novel interferon-γ release assay for the detection of SARS-CoV-2 specific T-cell response

**DOI:** 10.1101/2021.07.17.21260316

**Authors:** Daniela Huzly, Marcus Panning, Franziska Smely, Martin Enders, Johanna Komp, Daniel Steinmann

## Abstract

**Background:** The reliable detection of the T-cell mediated response to COVID-19 or COVID-19 vaccination is important for individual patient care and for monitoring the immune response e.g. in COVID-19 vaccine trials in a standardized fashion.

**Methods:** We used blood samples from health care workers (HCW) with or without history of COVID-19 to define test accuracy of a novel interferon-γ release assay. Usefulness of qualitative and quantitative results after COVID-19 vaccination was examined in HCW receiving homologous or heterologous vaccination regimens. For a real-life performance evaluation, we analysed interferon-γ response to complete vaccination in 149 patients receiving immunosuppressive or immune modulating therapies.

**Results:** Using a double-cut-off strategy integrating the result of background stimulation the assay had a specificity of 100%. Sensitivity of the IGRA was 83.5 and 100% in HCW after SARS-CoV-2 infection more or less than 6 months ago. Quantitative results showed significant differences between first and second vaccine dose, but no difference between homologous and heterologous vaccination regimen. The majority of immunocompromised patients showed no immune response or isolated T-cell or antibody response to complete vaccination.

**Conclusions:** The novel IGRA proved to be a highly specific and sensitive tool to detect the SARS-CoV-2 specific T-cell response to COVID-19 as well as COVID-19 vaccination. In perspective, it may serve as a standardized tool in COVID-19 vaccine trials and in clinical care of immunosuppressed patients.

## Introduction

The role of the T-cell immune response in SARS-CoV-2 infection is not entirely understood, but data from animal models show an important role for protection against coronavirus disease 2019 (COVID-19) mediated by CD4^+^ and CD8^+^ T cells and the production of interferon gamma (IFN-γ) [1, 2]. Since December 2020, the first vaccines against COVID-19 are available but the quality and duration of the immune response to vaccination remains unclear as of yet. Although numerous SARS-CoV-2 antibody detection assays were introduced with unprecedented speed and are now widely in use, the implementation of T cell assays lagged behind. However, besides analysing the antibody response, it will be of importance to investigate the T-cell mediated immune response, e.g. in vaccine trials or in a clinical setting for individual patient care. Therefore, easy to perform, validated, and ideally standardized T-cell assays are required [3].

Here we evaluated a novel commercially available IFN-γ release assay (IGRA) to analyse the SARS-CoV-2 specific T-cell response. IGRA have revolutionized the diagnosis of tuberculosis [4] and several other pathogens [5, 6]. In experimental settings, in-house SARS-CoV-2 IGRA were introduced recently [7]. The main advantage of this type of assay is the possibility to perform them without special equipment and with very short hands-on time. The IFN-γ results are expressed in international units and are therefore truly quantitative.

For our evaluation study, we asked immunocompetent healthcare workers (HCW) at Freiburg University Hospital to participate and offered measurement of SARS-CoV-2 antibodies. One group comprised HCW who had experienced mild SARS-CoV-2 infection in 2020 and another group had been vaccinated against COVID-19. For negative controls, we asked HCW without a history of COVID-19 prior to vaccination. After successful evaluation of the assay, we analysed the COVID-19 vaccine associated T-cell immune response in several patient groups including immunocompromised patients to evaluate the test performance in a real-life setting.

## Methods

### Study population

A total of 230 HCW agreed to participate. Of these, 107 had previous SARS-CoV-2 infection (COVID-19 group) proven by a positive SARS-CoV-2 RT-PCR and seroconversion. In the vaccine group, 38 were tested three weeks after the first and two weeks after the second dose of vaccination with BNT162b2 (Comirnaty®, BioNTech/Pfizer). Another 30 HCW were analysed three to four weeks after the first dose of vaccination with AZD1222 (Vaxzevria®, AstraZeneca). Of these 30 AZD1222 vacinees, 27 received a heterologous vaccination scheme with a second dose of mRNA-1273 (COVID-19 vaccine Moderna®, Moderna Biotech) and were sampled two weeks hereafter. Finally, 55 HCW were without history of SARS-CoV-2 infection or vaccination (no COVID-19 group). The Department of Occupational Medicine collected the HCW samples after informed consent. From all participants a serum sample was drawn for SARS-CoV-2 antibody testing together with a heparinized whole blood sample for the IGRA. Samples were tested independently without knowledge of SARS-CoV-2 antibody status or patient history. In the first three cases with blank control result above 100 mIU/ml, a second heparinized whole blood sample was drawn to exclude sampling error. For the real-life performance evaluation, all patient samples (n=149) sent to our routine diagnostic laboratory with the request for SARS-CoV-2 T-cell analysis and antibody detection after COVID-19 vaccination were included. Patients received various immunosuppressive or immune modulating therapies.

### SARS-CoV-2 interferon-γ release assay

The SARS-CoV-2 IGRA (EUROIMMUN, Lübeck, Germany) is based on the SARS-CoV-2 spike protein and was done according to the manufacturer’s instructions. A set of three tubes is required per whole blood sample: 1) SARS-CoV-2 IGRA BLANK without interferon activating substance to define the individual background stimulation. 2) SARS-CoV-2 IGRA TUBE containing antigens based on the S1 domain of SARS-CoV-2 spike protein. 3) SARS-CoV-2 IGRA STIM containing a mitogen for unspecific interferon stimulation for testing the viability and stimulation capacity of T cells and sufficient number of T cells in the whole blood samples. In brief, 500 µl of heparinized whole blood samples were used and incubated at 37 °C for 20 h to 24 h. After incubation, tubes were centrifuged at 8,000g for 10 min and 200 µl of supernatant was stored at 2-8 °C until measurement. For the IFN-γ enzyme-linked immunosorbent assay (ELISA), supernatant was diluted 1:5 with dilution buffer. Optical density (O.D.) was measured at 450 nm after enzymatic reaction. A set of six standards and two controls was used for each run. Samples with IFN-γ values above the standard curve were further diluted 1:10, 1:50 or 1:100, depending on the O.D. of the initial measurement. The manufacturer defined cut off is at >200 mIU/ml including a grey zone of 100 – 200 mIU/ml.

### Detection of SARS-CoV-2 antibodies

We used the following SARS-CoV-2 immunoglobuline G (IgG) assays to rule out past infection in the no COVID-19 and in the vaccine cohorts: Anti-SARS-CoV-2 ELISA IgG, detecting IgG antibodies against S1 (EUROIMMUN, Lübeck, Germany); recomWell SARS-CoV-2 IgG detecting IgG antibodies against the nucleoprotein (N) (Mikrogen, Munich, Germany); ADVIA Centaur SARS-CoV-2 total antibody (COV2T) assay, detecting total antibodies against S1-RBD (Siemens, Munich, Germany); and the Elecsys Anti-SARS-CoV-2, detecting total antibodies against N (Roche, Mannheim, Germany). Quantitative antibody levels as well as functional analysis of antibodies in the COVID-19 and vaccine groups are subject of a separate study.

### Statistical analysis

Data were analysed using IBM SPSS Statistics version 24 software (www.ibm.com/spss/statistics), GraphPad Prism version 9 (www.graphpad.com), and MedCalc version 19 (www.medcalc.com). We determined test accuracy, positive and negative predictive values and likelihood ratios assuming a disease prevalence of 10% and using only measured values of individuals after SARS-CoV-2 RT-PCR- and seroconversion-proven infection. For test accuracy calculation, we used two different approaches: we defined grey zone results as either positive or negative. As data had a non-Gaussian distribution, we used nonparametric tests throughout, a Mann-Whitney U test for single unpaired and Wilcoxon matched-pairs signed rank test for paired comparisons of medians. Receiver operating characteristic (ROC) analysis was used to define an optimum cut-off value at the Youden maximum index value. We calculated coefficient of variation for intra- and inter-assay variability. All raw data can be found in the supplementary file.

### Ethics

The study was approved by the ethics committee of Albert-Ludwigs University Freiburg (#20-1271, Nov 24, 2020 and #20-1271_1, Jan 18, 2021). Written informed consent was obtained from all participants.

## Results

We enrolled 230 HCW for the validation study. All HCW in the COVID-19 group had referred mild symptoms. Baseline characteristics of the HCW study group are shown in Table 1.

**Table 1.** Baseline characteristics of the 230 health care workers.

|  | COVID-19<br>>6<br>months<br>ago, n=91 | COVID-19<br><6<br>months<br>ago, n=16 | Vaccine group<br>BNT162b2,<br>n=38 | Vaccine<br>group<br>AZD1222,<br>n=30 | Vaccine<br>group<br>AZD1222/<br>mRNA-1273,<br>n=27 | No COVID-19 group,<br>n=55 |
| --- | --- | --- | --- | --- | --- | --- |
| Age in<br>years,<br>mean<br>(range) | 43.9 (22 -<br>64) | 38.1 (22 -<br>60) | 42.6 (19 - 63) | 44.2 (23 - 64) | 44.9 (23 -<br>64) | 42.58 (20 -<br>70) |
| Sex |  |  |  |  |  |  |
| Female % | 64.8 | 50 | 76.3 | 88.9 | 88.9 | 67.3 |
| Male % | 35.2 | 50 | 23.7 | 11.1 | 11.1 | 32.7 |

### IFN-γ concentrations in the no COVID-19 group

Out of 55 samples from the no COVID-19 group, 50 had IFN-γ concentrations below 100 mIU/ml, two between 100 and 200 mIU/ml, and three above 200 mIU/ml. Serum samples of these three individuals were reactive in several antibody assays against SARS-CoV-2 nucleoprotein (N) and spike protein (S1) (Table 2), suggesting past infection. Thus, we excluded these samples from further analysis. Serum samples from the 52 remaining individuals of this cohort tested negative with three different SARS-CoV-2 antibody assays and were therefore defined as truly negative controls. Specificity of the assay using 200 mIU/ml and 100 mIU/ml was 100% [95% confidence interval (CI) 93.2%-100%] and 96.2% (95% CI 86.8%-99.5%), respectively.

**Table 2:** SARS-CoV-2 antibody results of three IFN-y positive individuals of the “no COVID-19” group.

| Sample ID | S1-IgG ELISA,<br>EUROIMMUN | N-IgG ELISA,<br>Mikrogen | S1/RBD total AB,<br>Siemens | N total AB, Roche |
| --- | --- | --- | --- | --- |
| 94 | 0.435 (negative) | 26.8 (equivalent) | 4.45 (positive) | 22.2 (positive) |
| 95 | 1.2 (positive) | 5.73 (negative) | >10.00 (positive) | 2.4 (positive) |
| 100 | 1.52 (positive) | 14.7 (negative) | >10.00 (positive) | 27.7 (positive) |

Highest Youden index using ROC analysis was seen at 135 mIU/ml with a specificity of 98.1% (95% CI 89.8%-99.9%). We therefore re-analysed data using three different cut-offs with IFN-γ concentrations of 100 mIU/ml, 135 mIU/ml, and 200 mIU/ml.

### IFN-γ concentrations in HCW with previous SARS-CoV-2 infection (COVID-19 group)

A total of 107 samples from HCW with previous SARS-CoV-2 infection were available to us. In one case, the assay was invalid and thus excluded from the analysis. Of these, 90 HCW had blood samples drawn more than six months after RT-PCR proven SARS-CoV-2 infection. Eleven (12.2%) yielded IFN-γ concentrations below 100 mIU/ml, 14 (15.6%) between 100 and 200 mIU/ml, 13 (14.4%) between 135 and 200, and 65 (72.2%) above 200 mIU/ml. In this subgroup, we calculated a sensitivity of 71.4% defining IFN-γ concentrations 200 mIU/ml as true positive to detect SARS-CoV-2 infection in samples taken more than 6 months ago. Another 16 individuals were infected two to five months ago, 15 of which had IFN-γ concentrations above 200 mIU/ml and one was in our calculated as well as in the manufacturers grey-zone (151 mIU/ml). Sensitivity for recent infection in this subgroup was thus 93.8% or 100% defining grey-zone results as positive. Overall sensitivity to detect past infection using different cut-offs was 75.4%, 89.6% and 88.7% with cut-off 200 mIU/ml, 100 mIU/ml or 135 mIU/ml, respectively.

### Background IFN-γ concentrations and adapted performance calculation

In the initial validation, BLANK values were below 50 mIU/ml (mean 35.2 mIU/ml, range 0-1520 mIU/ml) in most patients, but 23/230 (10%) showed elevated IFN-γ concentrations above 100 mIU/ml including the two false positive samples (see above). We therefore aimed to include the BLANK values into the result interpretation and to circumvent the need of a grey-zone. Subsequently, we used a cut-off of equal or above 135 mIU/ml in individuals with background stimulation below 100 mIU/ml and a cut-off of 200 mIU/ml in individuals with higher background stimulation. Using our BLANK value adapted approach the specificity reached 100 % and overall sensitivity to detect past infection was 86.8% (Table 3). In detail, it was 100% for infection less than five months ago and 83.5% for infection more than 6 months ago. Predictive values and accuracy of the different cut-off strategies are shown in Table 3. Of note, 20 out of 107 (18.7%) HCW several months after SARS-CoV-2 infection were negative for SARS-CoV-2 S1-IgG but tested positive for IFN-γ stimulation using the BLANK value adapted cut-off. Three of five HCW who were non-reactive even in the highly sensitive total-antibody assays were positive showing IFN-γ concentrations between 136 and 630 mIU/ml. In nine of 11 cases with equivocal SARS-CoV-2 S1-IgG, IFN-γ results were positive.

**Table 3:** Diagnostic test accuracy of the EUROIMMUN interferon-γ release assay using different cut-offs at an estimated COVID-19 prevalence of 10%.

| | Cut-off >100 mIU/ml | Cut-off ≥135 mIU/ml | Adapted cut-off* |
| --- | --- | --- | --- |
| Sensitivity % (95% CI) | 89.6 (82.2 - 94.7) | 88.7 (81.1 - 94.0) | 86.8 (78.8 - 92.6) |
| Specificity | 96.2 (86.8 - 99.5) | 98.1 (89.7 - 99.9) | 100 (93.2 - 100) |
| Negative predictive value | 98.8 (97.9 - 99.3) | 98.7 (97.9 - 99.3) | 98.6 (97.7 - 99.1) |
| Positive predictive value | 72.2 (39.9 - 90.9) | 83.7 (42.4 - 97.3) | 100 |
| Accuracy | 95.5 (90.9 - 98.2) | 97.1 (93.2 - 99.1) | 98.7 (95.4 - 99.8) |

\*Adapted cut-off: Cut-off ≥ 135 mIU/ml if BLANK < 100 mIU/ml and ≥ 200mIU/ml if BLANK ≥ 100 mIU/ml

### IFN-γ concentrations in individuals after COVID-19 vaccination

Next, we analysed IFN-γ concentrations in HCW after COVID-19 vaccination. In the BNT162b2 group, 36/38 (95%) had IFN-γ concentrations above 200 mIU/ml after the first dose and 38/38 (100%) after the second dose, respectively (Figure 1). Of note, two participants had no IFN-γ release after the first vaccination (6 and 9 mIU/ml), but subsequently reacted after the second dose of BNT162b2. In individuals receiving AZD1222, 28/30 (93%) had IFN-γ concentrations above 200 after the first dose, one had 190 mIU/ml and one did not show a specific IFN-γ release (64 mIU/ml). All 27 HCW who were vaccinated with the Moderna vaccine mRNA1273 after receiving a first dose with AZD1222 had IFN-γ concentrations clearly above 200 mIU/ml.

**Figure 1:**
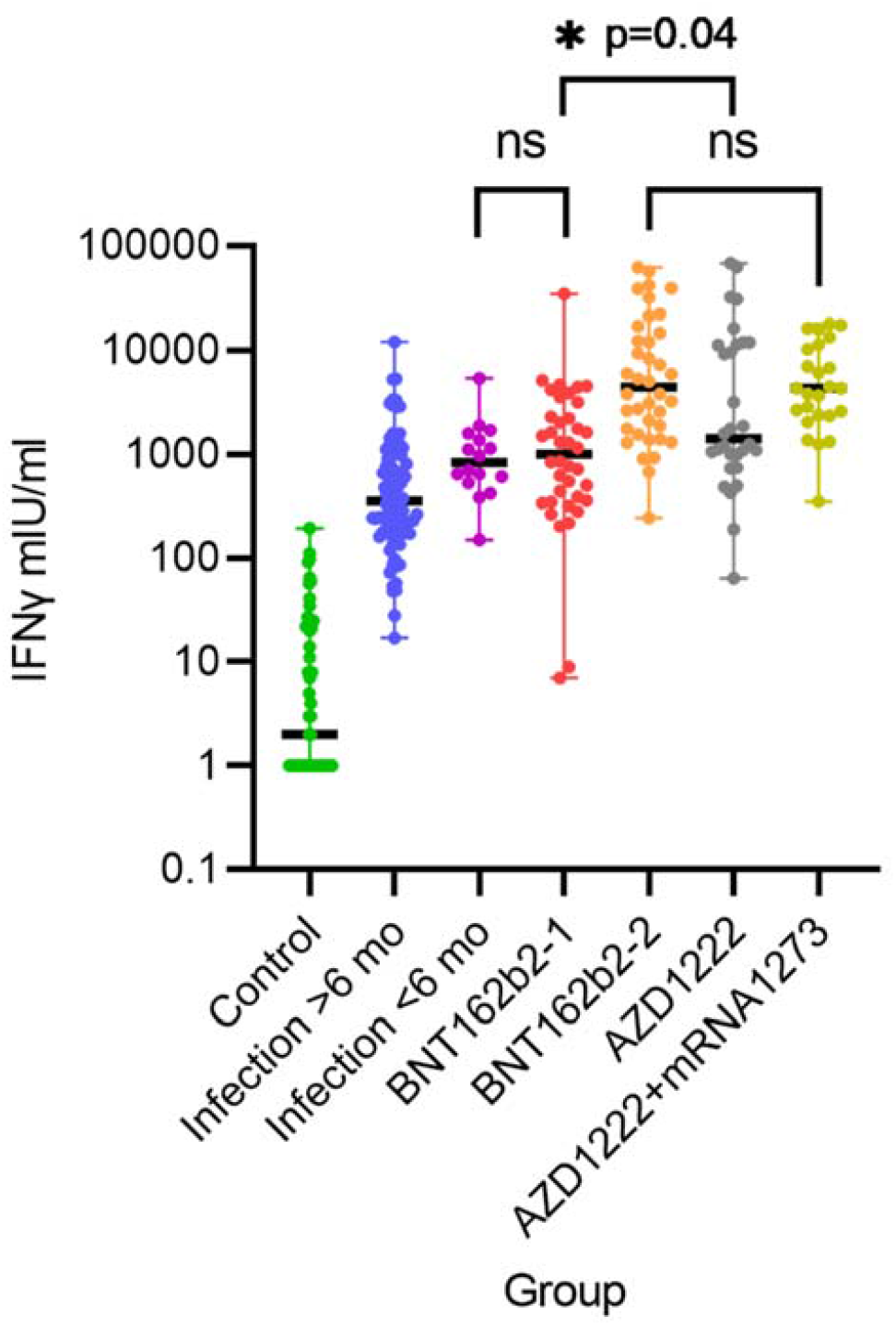
IFN-y release after stimulation with SARS-CoV-2 spike antigen in different groups. Differences are highly significant with p<0.001 unless indicated. BNT162b2-1 and -2 denotes health care workers (HCW) after the first and second dose of BNT162b2 (n=38), AZD1222 denotes HCW after first dose of AZD1222 (n=30) and AZD1222+mRNA1273 after the second dose with mRNA-1273 (heterologous vaccination scheme, n=27). ns= not significant

### Comparison of IFN-γ concentrations in COVID-19 patients versus vaccinated HCW

Median IFN-γ concentration was significantly different between individuals with past infection more than 6 months vs. less than 6 months ago (Mann-Whitney U test, p=0.001) (Figure 1). IFN-γ concentrations after first dose of BNT162b2 vaccine were similar as those after recent infection and significantly lower than concentrations after the second dose (Wilcoxon test, p=0.0010). Median IFN-γ concentration after the first dose of AZD1222 was higher than after the first dose of BNT162B2 with a large range (Mann-Whitney U test, p=0.04). Median concentration after heterologous booster immunization with mRNA1273 was similar to median concentration after second dose with BNT162b2. Mean, median, standard deviation and range are shown in Table 4. In the HCW group with past infection more than 6 months ago median IFN-γ concentration was significantly higher in individuals older than 50 years than in younger individuals (Mann-Whitney U test, p<0.0001). There was no significant difference between age groups in vaccinated HCW or in those who had experienced the infection less than 6 months ago.

**Table 4:** Interferon-γ results in mIU/ml in the different study groups.

| Study group | n | mean | SD* | median | min | max |
| --- | --- | --- | --- | --- | --- | --- |
| No COVID-19 | 52 | 20.37 | 37.8 | 2.0 | 0 | 193 |
| Past infection >6mo | 90 | 885.1 | 1584.1 | 355.5 | 17 | 12149 |
| Past infection <6mo | 16 | 1205.9 | 1224.3 | 837.5 | 151 | 5383 |
| BNT162b2 1. dose | 38 | 2444.6 | 5522.4 | 1007.5 | 7 | 34300 |
| BNT162b2 2. dose | 38 | 14268.6 | 18437.0 | 5197.5 | 245 | 67049 |
| AZD1222 1. dose | 30 | 9847.1 | 17207.0 | 1421.0 | 64 | 67748 |
| AZD1222/mRNA1273 | 27 | 6492.0 | 16969.2 | 4440.0 | 245 | 67049 |

\*SD Satn

### IFN-y response in immunocompromised patients

After validation and implementation of the assay in our routine diagnostic lab, we encouraged colleagues to analyse the immune response in immunocompromised patients after complete COVID-19 vaccination. These patients (n=149) had various underlying diseases and received immunosuppressive or immune modulating therapies or had variable immunodeficiency (Table 5). The majority of immunocompromised patients showed an inadequate immune response to vaccination, which was in most cases done using BNT162b2 and in few cases applying the heterologous vaccination scheme AZD1222/BNT162b2. Of note, most responding patients had isolated T-cell or antibody response (Table 5). 13 of 20 patients who had received B-cell depleting therapy up to 12 months ago had isolated T-cell response with IFN-γ release >1000 mIU/ml.

**Table 5.** Combined immune response (detection of SARS-CoV-2 S1-IgG and/or IFN-y response) to vaccination in immunocompromised patients and healthy adults.

| Underlying disease | S1-IgG negative AND IGRA negative or invalid<br>N (%) | IGRA positive, S1-IgG negative<br>N (%) | S1-IgG positive, IGRA negative or invalid<br>N (%) | IGRA and S1-IgG positive<br>N (%) | IGRA invalid<br>N (%) |
| --- | --- | --- | --- | --- | --- |
| Lung transplant (n=33) | 29 (87,9) | 1 (3.0) | 2 (6.0) | 1 (3.0) | 8 (24.2) |
| Kidney transplant (n=24) | 14 (58.3) | 3 (12.5) | 5 (20.8) | 2 (8.3) | 9 (37.5) |
| Haemato-oncological (n=24) | 12 (50) | 6 (25.0) | 2 (8.3) | 4 (16.6) | 3 (12.5) |
| Stem cell transplant (n=19) | 3 (15.8) | 3 (15.8) | 6 (31.6) | 7 (36.8) | 3 (15.8) |
| Autoimmune disorder (n=35) | 7 (20.0) | 14 (40.0) | 4 (11.4) | 10 (28.6) | 2 (5.7) |
| Variable immunodeficiency (n=14) | 6 (42.9) | 1 (7.1) | 1 (7.1) | 6 (42.8) | 0 |
| HCW BNT162b2 (n=38) | 0 | 0 | 0 | 38 (100) | 0 |
| HCW AZD1222/mRNA1273 (n=27) | 0 | 0 | 0 | 27 (100) | 0 |

## Discussion

In this study, the EUROIMMUN SARS-CoV-2 IGRA demonstrated a cut-off dependent specificity of 96.3-100% and sensitivity of 75.4-89.6%. We used a rigorously tested set of samples to evaluate the IGRA thoroughly. The IGRA proved to be a sensitive and specific tool for detecting the cellular immune response to COVID-19 or COVID-19 vaccination. Technically, it is easy to perform and can be rapidly implemented in routine diagnostic laboratories. This will assist e.g. in patient care and future COVID-19 vaccination trials as it will allow on site testing of the cellular immune response using standardized and widely available assays.

Understanding the immune response to COVID-19 is essential for the development of preventive strategies against COVID-19. Since the end of 2020 several vaccines have been authorized by medicines agencies worldwide and mass vaccination campaigns in some countries have been very successful in reducing case numbers significantly [8, 9]. Nevertheless, breakthrough infections have been reported to occur with all vaccines and it will be important to define the correlates of protection after vaccination as well as after infection [10-14]. Serological assays to quantify SARS-CoV-2 specific antibodies against spike protein (S1) or the receptor binding domain as well as surrogate neutralization assays for the detection of protective antibodies have been described [15, 16]. However, from recent communications of breakthrough infections in healthy and immunosuppressed adults it appears that the humoral response alone may not be sufficient for protection against infection and disease [11, 14, 17]. Longitudinal studies of COVID-19 patients have observed a significant association of a specific T cell response with milder disease in the absence of SARS-CoV-2 IgG seroconversion and even in agammaglobulinaemic and B-cell depleted patients suggesting that T cell responses may be important for control of SARS-CoV-2 infection [18-24]. Of note, T cell-responses in convalescents and after COVID-19 vaccination seem not to be affected by mutations found in SARS-CoV-2 variants of concern [25-28], whereas neutralizing capacity of antibodies is significantly reduced [29]. It is therefore of importance to measure T-cell mediated reactivity to evaluate immune response to vaccination.

For multicentre vaccination studies standardized and easy to perform assays to measure T cell response in large scale manner are required. IFN-γ is a key cytokine for T cell-mediated immune response to specific antigens [30, 31]. The expression of IFN-γ can therefore be used as a marker of pathogen-specific immunity. There are different ways to measure IFNγ release *ex vivo*, but for most assays, different steps for separation of lymphocytes, cell count and staining are necessary. If measurement at once is not feasible, specific freezing protocols are required. The ELISA-based IGRA is comparably easy to perform. Whole blood is transferred to the stimulation tubes and the separated plasma can be stored at 2-8 °C up to 4 weeks and at -20 °C for at least three months until measurement. Of note, the results of different assay types for IFN-γ release upon stimulation are difficult to compare [30] and there is no true gold standard for the measurement of cellular immune response. Thus, for diagnostic test accuracy evaluation other information has to be used [32]. We used samples from HCW without history of COVID-19 or COVID-19 vaccination and without detectable SARS-CoV-2 antibodies using various antibody detection formats for specificity analysis. Several authors have observed SARS-CoV-2 reactive T cells in negative cohorts [33, 34]. The here evaluated IGRA was negative in all 52 healthy controls in our study using the adapted cut-off strategy, showing very high specificity, which is an essential requirement for its suitability for large-scale studies. It further supports the notion that the assay dos not interfere with pre-existing and cross-reactive T cells from previous common coronavirus infections.

Diagnosis of past infection without history of PCR positivity is difficult using SARS-CoV-2 antibody-based testing alone, as antibodies wane after several months and in some cases become undetectable even with the most sensitive antibody assays [16]. Highly specific T-cell assays may help proving specificity of low positive or grey-zone antibody results. In a recent study with COVID-19 convalescents, a cellular immune response had been detectable in 78% of non-seroconverting and 80% of seroconverting patients by ELISPOT using different peptide pools and proteins. The authors found highest specificity but lowest sensitivity using the S1 protein [18]. The EUROIMMUN IGRA uses only an S1 peptide pool for stimulation; however, using the adapted cut-off specific IFN-y release was detected in 100% and 83.5% of individuals after infection less and more than 6 months ago, respectively.

For the use in vaccination studies quantification of the T-cell response must be reliable. We were able to show significant differences between the first and the second vaccination among groups as well as individuals and between different vaccines. Similar results have been reported recently in a study using the same IGRA [35]. Reproducibility was good and the assay showed high tolerance against variations of standard procedures (data not shown). These properties make the assay highly suitable for routine diagnostic laboratories and opens up possibilities to analyze the immune response beyond using antibody detection assays in vaccinated patients especially under immunosuppressing or immune modulating medications. Importantly, we were able to show that B-cell depleted patients in most cases mount a strong T-cell response to COVID-19 vaccination and are thus probably protected against severe disease [24]. However, this was not seen in patients after organ transplantation confirming previous studies [36]. These patients can only be protected by vaccination of their close contacts and non-pharmaceutical intervention strategies.

Limitation for the use of such a biological assay is the functionality of the T-cell response. Even low-dose steroid treatment may interfere with IGRA testing, as has been shown for Quantiferon TB in immunocompetent children (34). We have seen in one of our patients that even short-term steroid treatment may be of relevance. The patient yielded a negative mitogen control and a negative response to SARS-CoV-2 antigen (46 and 15 mIU/ml) three weeks after vector/mRNA heterologous vaccination, but SARS-CoV-2 S1-IgG was detectable. She was on high dose steroids when blood was drawn, but not during vaccination. After stopping steroid treatment, we repeated the assay and detected 985 mIU/ml after antigen stimulation with high positive mitogen control (1880 mIU/ml). Further, the interpretation of grey-zone results is always difficult, so we developed a double cut-off strategy including BLANK results in result interpretation. Diagnostic test accuracy analysis showed best accuracy with similar negative predictive values using our adopted cut-off strategy.

In conclusion, the EUROIMMUN IGRA is an easy to perform assay for the detection of specific T-cell response against SARS-CoV-2 spike antigen. The assay is highly specific and sensitive. During a performance evaluation of the assay we were able to show that patients on immunosuppressive regimens may have isolated T-cell or antibody response or in several cases do not respond at all. For clinical practice, we recommend asking for steroid treatment before performing such an assay and if possible pausing treatment before drawing blood. We propose that measurement of immune response to vaccination should always include an analysis of the T-cell response and the EUROIMMUN IGRA proved to be highly suitable for this purpose.

## Data Availability

All raw data can be found in the supplementary file.

## Acknowledgement

We are grateful to Ingeborg Hanselmann for expert technical assistance. We would like to thank all health care workers and patients for their participation and support.

