## Supplemental Table 1 for "Validation and performance evaluation of a novel interferon-γ release assay for the detection of SARS-CoV-2 specific T-cell response"

Supplemental Tables 1a-d. TUBE = S1 antigen stimulation; BLANK = no antigen, background stimulation; STIM = mitogen control. BLANK is subtracted from TUBE or STIM for result calculation.

Table 1a. Results of IFNγ release in 55 health care workers without history of SARS-CoV2 infection (NCD = 3 HCWs with diagnosed past infection)

| **Participant** | **Category** | **TUBE** | **BLANK** | **STIM** | **TUBE result** | **STIM result** |
| --- | --- | --- | --- | --- | --- | --- |
| 2 | NK | 2 | 2 | 1445 | 0 | 1443 |
| 29 | NK | 96 | 1 | 1125 | 95 | 1124 |
| 43 | NK | 0 | 0 | 870 | 0 | 870 |
| 49 | NK | 21 | 1 | 1550 | 20 | 1549 |
| 82 | NK | 1 | 1 | 1595 | 0 | 1594 |
| 83 | NK | 25 | 18 | 1595 | 7 | 1577 |
| 84 | NK | 95 | 93 | 1595 | 2 | 1502 |
| 85 | NK | 1 | 1 | 1595 | 0 | 1594 |
| 86 | NK | 9 | 1 | 1595 | 8 | 1594 |
| 87 | NK | 2 | 1 | 1595 | 1 | 1594 |
| 88 | NK | 1 | 1 | 1595 | 0 | 1594 |
| 89 | NK | 127 | 127 | 1595 | 0 | 1468 |
| 90 | NK | 131 | 96 | 1595 | 35 | 1499 |
| 91 | NK | 4 | 1 | 1595 | 3 | 1594 |
| 92 | NK | 91 | 80 | 1595 | 11 | 1515 |
| 93 | NK | 28 | 1 | 1595 | 27 | 1594 |
| 96 | NK | 324 | 131 | 1595 | 193 | 1464 |
| 97 | NK | 10 | 2 | 1595 | 8 | 1593 |
| 98 | NK | 121 | 100 | 1595 | 21 | 1495 |
| 99 | NK | 1 | 1 | 1595 | 0 | 1594 |
| 101 | NK | 2 | 1 | 1595 | 1 | 1594 |
| 103 | NK | 96 | 4 | 1595 | 92 | 1591 |
| 104 | NK | 1 | 1 | 1595 | 0 | 1594 |
| 105 | NK | 11 | 11 | 1595 | 0 | 1584 |
| 106 | NK | 2 | 1 | 1595 | 1 | 1594 |
| 107 | NK | 62 | 5 | 1595 | 57 | 1590 |
| 109 | NK | 10 | 7 | 1595 | 3 | 1588 |
| 110 | NK | 5 | 1 | 1595 | 4 | 1594 |
| 111 | NK | 23 | 1 | 1595 | 22 | 1594 |
| 112 | NK | 99 | 1 | 1595 | 98 | 1594 |
| 113 | NK | 250 | 139 | 1595 | 111 | 1456 |
| 115 | NK | 53 | 12 | 1595 | 41 | 1583 |
| 116 | NK | 11 | 10 | 1595 | 1 | 1585 |
| 117 | NK | 1 | 1 | 1595 | 0 | 1594 |
| 118 | NK | 3 | 3 | 1595 | 0 | 1592 |
| 119 | NK | 2 | 1 | 1595 | 1 | 1594 |
| 120 | NK | 376 | 351 | 1595 | 25 | 1244 |
| 121 | NK | 94 | 29 | 1595 | 65 | 1566 |
| 125 | NK | 2 | 2 | 1595 | 0 | 1593 |
| 127 | NK | 1 | 1 | 1595 | 0 | 1594 |
| 141 | NK | 15 | 1 | 1595 | 14 | 1594 |
| 142 | NK | 2 | 1 | 1595 | 1 | 1594 |
| 143 | NK | 1 | 1 | 1595 | 0 | 1594 |
| 144 | NK | 64 | 4 | 1370 | 60 | 1366 |
| 145 | NK | 1 | 1 | 1555 | 0 | 1554 |
| 146 | NK | 1 | 1 | 1595 | 0 | 1594 |
| 147 | NK | 2 | 1 | 1595 | 1 | 1594 |
| 151 | NK | 3 | 1 | 1595 | 2 | 1594 |
| 180 | NK | 76 | 53 | 1595 | 23 | 1542 |
| 183 | NK | 6 | 1 | 1595 | 5 | 1594 |
| 191 | NK | 1 | 1 | 220 | 0 | 219 |
| 215 | NK | 1 | 1 | 1095 | 0 | 1094 |
| 94 | NCD | 352 | 1 | 1595 | 351 | 1594 |
| 95 | NCD | 214 | 7 | 1595 | 207 | 1588 |
| 100 | NCD | 875 | 95 | 1595 | 780 | 1500 |

Table 1b. Results of IFNγ release in 107 health care workers with history of SARS-CoV2 infection >6mo ago (INF) or <6ml ago (INF2). *Participant #6 was excluded from further analysis

| **Participant** | **Category** | **TUBE** | **BLANK** | **STIM** | **TUBE result** | **STIM result** |
| --- | --- | --- | --- | --- | --- | --- |
| 1 | INF | 388 | 10 | 1445 | 378 | 1435 |
| 3 | INF | 102 | 14 | 1250 | 88 | 1236 |
| 4 | INF | 293 | 1 | 1595 | 292 | 1594 |
| 5 | INF | 249 | 1 | 1310 | 248 | 1309 |
| 6* | INF | 1520 | 1520 | 1595 |  | 75 |
| 7 | INF | 495 | 1 | 630 | 494 | 629 |
| 8 | INF | 29 | 1 | 1595 | 28 | 1594 |
| 9 | INF | 631 | 1 | 1595 | 630 | 1594 |
| 10 | INF | 181 | 1 | 740 | 180 | 739 |
| 11 | INF | 55 | 1 | 1595 | 54 | 1594 |
| 12 | INF | 151 | 134 | 1340 | 17 | 1206 |
| 13 | INF | 490 | 262 | 840 | 228 | 578 |
| 14 | INF | 478 | 104 | 625 | 374 | 521 |
| 15 | INF | 750 | 18 | 1595 | 732 | 1577 |
| 16 | INF | 560 | 63 | 1595 | 497 | 1532 |
| 17 | INF | 262 | 42 | 285 | 220 | 243 |
| 18 | INF | 314 | 67 | 630 | 247 | 563 |
| 19 | INF | 1331 | 86 | 1595 | 1245 | 1509 |
| 20 | INF | 811 | 1 | 1595 | 810 | 1594 |
| 21 | INF | 911 | 1 | 1595 | 910 | 1594 |
| 22 | INF | 3365 | 2 | 1595 | 3363 | 1593 |
| 23 | INF | 12150 | 1 | 1595 | 12149 | 1594 |
| 24 | INF | 231 | 1 | 1370 | 230 | 1369 |
| 25 | INF | 1595 | 166 | 1250 | 1429 | 1084 |
| 26 | INF | 1540 | 94 | 1595 | 1446 | 1501 |
| 27 | INF | 386 | 1 | 1500 | 385 | 1499 |
| 28 | INF | 99 | 1 | 885 | 98 | 884 |
| 30 | INF | 178 | 1 | 1595 | 177 | 1594 |
| 31 | INF | 1265 | 106 | 311 | 1159 | 205 |
| 32 | INF | 600 | 190 | 1595 | 410 | 1405 |
| 33 | INF | 348 | 15 | 1595 | 333 | 1580 |
| 34 | INF | 116 | 43 | 545 | 73 | 502 |
| 35 | INF | 86 | 2 | 1280 | 84 | 1278 |
| 36 | INF | 420 | 2 | 1230 | 418 | 1228 |
| 37 | INF | 141 | 5 | 525 | 136 | 520 |
| 38 | INF | 241 | 1 | 1185 | 240 | 1184 |
| 39 | INF | 3116 | 1 | 1595 | 3115 | 1594 |
| 40 | INF | 227 | 3 | 1553 | 224 | 1550 |
| 41 | INF | 226 | 1 | 1595 | 225 | 1594 |
| 42 | INF | 228 | 1 | 1595 | 227 | 1594 |
| 44 | INF | 181 | 5 | 1595 | 176 | 1590 |
| 45 | INF | 267 | 1 | 1595 | 266 | 1594 |
| 46 | INF | 706 | 95 | 1330 | 611 | 1235 |
| 47 | INF | 303 | 5 | 1595 | 298 | 1590 |
| 48 | INF | 2891 | 38 | 1595 | 2853 | 1557 |
| 51 | INF | 289 | 1 | 1595 | 288 | 1594 |
| 52 | INF | 181 | 6 | 510 | 175 | 504 |
| 53 | INF | 177 | 5 | 415 | 172 | 410 |
| 54 | INF | 1596 | 379 | 1595 | 1217 | 1216 |
| 55 | INF | 745 | 1 | 1595 | 744 | 1594 |
| 56 | INF | 225 | 1 | 1045 | 224 | 1044 |
| 57 | INF | 1595 | 2 | 1345 | 1593 | 1343 |
| 58 | INF | 230 | 1 | 1595 | 229 | 1594 |
| 59 | INF | 484 | 2 | 1125 | 482 | 1123 |
| 60 | INF | 468 | 307 | 1595 | 161 | 1288 |
| 61 | INF | 246 | 2 | 1545 | 244 | 1543 |
| 62 | INF | 221 | 102 | 1595 | 119 | 1493 |
| 63 | INF | 1055 | 2 | 1595 | 1053 | 1593 |
| 64 | INF | 3350 | 1 | 1595 | 3349 | 1594 |
| 65 | INF | 595 | 2 | 1595 | 593 | 1593 |
| 66 | INF | 5301 | 1 | 1010 | 5300 | 1009 |
| 67 | INF | 670 | 3 | 1595 | 667 | 1592 |
| 68 | INF | 1595 | 1 | 1595 | 1594 | 1594 |
| 69 | INF | 2887 | 2 | 1595 | 2885 | 1593 |
| 70 | INF | 159 | 1 | 1595 | 158 | 1594 |
| 71 | INF | 297 | 1 | 1595 | 296 | 1594 |
| 72 | INF | 1385 | 6 | 1595 | 1379 | 1589 |
| 73 | INF | 545 | 5 | 1595 | 540 | 1590 |
| 74 | INF | 3000 | 23 | 1595 | 2977 | 1572 |
| 75 | INF | 1211 | 1 | 1595 | 1210 | 1594 |
| 76 | INF | 685 | 18 | 1595 | 667 | 1577 |
| 78 | INF | 766 | 1 | 1595 | 765 | 1594 |
| 79 | INF | 1111 | 1 | 1505 | 1110 | 1504 |
| 80 | INF | 670 | 1 | 1595 | 669 | 1594 |
| 102 | INF | 179 | 3 | 1595 | 176 | 1592 |
| 108 | INF | 194 | 146 | 1595 | 48 | 1449 |
| 122 | INF | 66 | 17 | 1595 | 49 | 1578 |
| 123 | INF | 1351 | 401 | 1595 | 950 | 1194 |
| 148 | INF | 146 | 1 | 1595 | 145 | 1594 |
| 149 | INF | 483 | 111 | 1595 | 372 | 1484 |
| 150 | INF | 403 | 64 | 1595 | 339 | 1531 |
| 152 | INF | 196 | 1 | 1595 | 195 | 1594 |
| 164 | INF | 1166 | 99 | 1595 | 1067 | 1496 |
| 167 | INF | 58 | 1 | 1595 | 57 | 1594 |
| 181 | INF | 986 | 1 | 1595 | 985 | 1594 |
| 184 | INF | 136 | 1 | 1595 | 135 | 1594 |
| 188 | INF | 293 | 29 | 1595 | 264 | 1566 |
| 196 | INF | 935 | 320 | 1210 | 615 | 890 |
| 197 | INF | 216 | 129 | 388 | 87 | 259 |
| 239 | INF | 296 | 123 | 1415 | 173 | 1292 |
| 276 | INF | 5350 | 2 | 1865 | 5348 | 1863 |
| 50 | INF2 | 1135 | 14 | 1595 | 1121 | 1581 |
| 77 | INF2 | 182 | 31 | 1595 | 151 | 1564 |
| 81 | INF2 | 1150 | 11 | 1595 | 1139 | 1584 |
| 114 | INF2 | 425 | 1 | 1595 | 424 | 1594 |
| 163 | INF2 | 695 | 42 | 1595 | 653 | 1553 |
| 166 | INF2 | 1896 | 1 | 1595 | 1895 | 1594 |
| 172 | INF2 | 976 | 1 | 1595 | 975 | 1594 |
| 182 | INF2 | 1596 | 1 | 1595 | 1595 | 1594 |
| 194 | INF2 | 701 | 1 | 1595 | 700 | 1594 |
| 195 | INF2 | 5400 | 17 | 1595 | 5383 | 1578 |
| 198 | INF2 | 580 | 48 | 945 | 532 | 897 |
| 228 | INF2 | 616 | 1 | 1595 | 615 | 1594 |
| 243 | INF2 | 1368 | 1 | 1595 | 1367 | 1594 |
| 274 | INF2 | 396 | 9 | 1970 | 387 | 1961 |
| 277 | INF2 | 655 | 4 | 1970 | 651 | 1966 |
| 282 | INF2 | 1710 | 3 | 2000 | 1707 | 1997 |

Table 1c. Results of IFNγ release in 38 health care workers after the first (VB1) and second (VB2) immunization with BNT162b2

| **Participant** | **Category** | **TUBE** | **BLANK** | **STIM** | **TUBE result** | **STIM result** |
| --- | --- | --- | --- | --- | --- | --- |
| 124 | VB1 | 348 | 1 | 1595 | 347 | 1594 |
| 126 | VB1 | 3900 | 2 | 1595 | 3898 | 1593 |
| 128 | VB1 | 4455 | 16 | 1595 | 4439 | 1579 |
| 129 | VB1 | 4311 | 96 | 1595 | 4215 | 1499 |
| 130 | VB1 | 334 | 68 | 1060 | 266 | 992 |
| 131 | VB1 | 206 | 2 | 1055 | 204 | 1053 |
| 132 | VB1 | 1315 | 1 | 1110 | 1314 | 1109 |
| 133 | VB1 | 1645 | 7 | 1595 | 1638 | 1588 |
| 134 | VB1 | 10 | 3 | 1595 | 7 | 1592 |
| 135 | VB1 | 10 | 1 | 1595 | 9 | 1594 |
| 136 | VB1 | 219 | 1 | 1595 | 218 | 1594 |
| 137 | VB1 | 34301 | 1 | 1595 | 34300 | 1594 |
| 138 | VB1 | 4550 | 30 | 1595 | 4520 | 1565 |
| 139 | VB1 | 506 | 1 | 1595 | 505 | 1594 |
| 140 | VB1 | 2030 | 1 | 1595 | 2029 | 1594 |
| 154 | VB1 | 5150 | 10 | 1595 | 5140 | 1585 |
| 155 | VB1 | 1760 | 1 | 1595 | 1759 | 1594 |
| 156 | VB1 | 2295 | 1 | 1170 | 2294 | 1169 |
| 157 | VB1 | 2240 | 20 | 1595 | 2220 | 1575 |
| 158 | VB1 | 626 | 1 | 1595 | 625 | 1594 |
| 159 | VB1 | 1395 | 106 | 1595 | 1289 | 1489 |
| 160 | VB1 | 1170 | 15 | 1595 | 1155 | 1580 |
| 161 | VB1 | 725 | 3 | 1595 | 722 | 1592 |
| 162 | VB1 | 1625 | 1 | 1595 | 1624 | 1594 |
| 165 | VB1 | 1510 | 3 | 1595 | 1507 | 1592 |
| 173 | VB1 | 391 | 1 | 1595 | 390 | 1594 |
| 174 | VB1 | 556 | 1 | 1335 | 555 | 1334 |
| 175 | VB1 | 280 | 1 | 1595 | 279 | 1594 |
| 176 | VB1 | 861 | 1 | 1000 | 860 | 999 |
| 177 | VB1 | 446 | 1 | 840 | 445 | 839 |
| 178 | VB1 | 3581 | 1 | 1595 | 3580 | 1594 |
| 187 | VB1 | 369 | 6 | 1595 | 363 | 1589 |
| 189 | VB1 | 319 | 1 | 430 | 318 | 429 |
| 193 | VB1 | 4716 | 1 | 497 | 4715 | 496 |
| 204 | VB1 | 3181 | 1 | 1595 | 3180 | 1594 |
| 225 | VB1 | 348 | 7 | 1595 | 341 | 1588 |
| 226 | VB1 | 791 | 19 | 1595 | 772 | 1576 |
| 227 | VB1 | 1000 | 149 | 1595 | 851 | 1446 |
| 131 | VB2 | 928 | 1 | 1885 | 927 | 1884 |
| 130 | VB2 | 1435 | 36 | 1885 | 1399 | 1849 |
| 129 | VB2 | 14700 | 1 | 1885 | 14699 | 1884 |
| 161 | VB2 | 6000 | 2 | 1885 | 5998 | 1883 |
| 138 | VB2 | 5351 | 1 | 1885 | 5350 | 1884 |
| 132 | VB2 | 3836 | 1 | 1110 | 3835 | 1109 |
| 156 | VB2 | 2750 | 1 | 1885 | 2749 | 1884 |
| 137 | VB2 | 12401 | 1 | 1885 | 12400 | 1884 |
| 136 | VB2 | 246 | 1 | 1885 | 245 | 1884 |
| 154 | VB2 | 41805 | 20 | 1885 | 41785 | 1865 |
| 134 | VB2 | 9350 | 1 | 1885 | 9349 | 1884 |
| 135 | VB2 | 911 | 1 | 1885 | 910 | 1884 |
| 155 | VB2 | 8350 | 9 | 1885 | 8341 | 1876 |
| 126 | VB2 | 3111 | 1 | 1885 | 3110 | 1884 |
| 124 | VB2 | 1895 | 1 | 1885 | 1894 | 1884 |
| 139 | VB2 | 21151 | 1 | 1885 | 21150 | 1884 |
| 140 | VB2 | 62252 | 2 | 1885 | 62250 | 1883 |
| 165 | VB2 | 3851 | 19 | 1885 | 3832 | 1866 |
| 162 | VB2 | 7251 | 1 | 1885 | 7250 | 1884 |
| 157 | VB2 | 22300 | 8 | 1885 | 22292 | 1877 |
| 159 | VB2 | 1595 | 35 | 1885 | 1560 | 1850 |
| 158 | VB2 | 680 | 1 | 1885 | 679 | 1884 |
| 133 | VB2 | 5050 | 5 | 1885 | 5045 | 1880 |
| 160 | VB2 | 12100 | 4 | 1885 | 12096 | 1881 |
| 128 | VB2 | 46800 | 69 | 1885 | 46731 | 1816 |
| 187 | VB2 | 53100 | 8 | 1885 | 53092 | 1877 |
| 175 | VB2 | 36901 | 1 | 1885 | 36900 | 1884 |
| 173 | VB2 | 1296 | 1 | 1885 | 1295 | 1884 |
| 176 | VB2 | 1306 | 1 | 1000 | 1305 | 999 |
| 177 | VB2 | 1391 | 1 | 840 | 1390 | 839 |
| 174 | VB2 | 22501 | 1 | 1310 | 22500 | 1309 |
| 178 | VB2 | 67050 | 1 | 1885 | 67049 | 1884 |
| 204 | VB2 | 2591 | 1 | 1885 | 2590 | 1884 |
| 227 | VB2 | 2063 | 295 | 1885 | 1768 | 1590 |
| 225 | VB2 | 29250 | 65 | 1885 | 29185 | 1820 |
| 226 | VB2 | 25251 | 75 | 1885 | 25176 | 1810 |
| 193 | VB2 | 1446 | 1 | 1885 | 1445 | 1884 |
| 189 | VB2 | 2635 | 1 | 430 | 2634 | 429 |

Table 1d. Results of IFNγ release in 30 health care workers after immunization with one dose of AZD1222 (VA) and in 27/30 after a heterologous dose of mRNA1273 (VAM)

| **Participant** | **Category** | **TUBE** | **BLANK** | **STIM** | **TUBE result** | **STIM result** |
| --- | --- | --- | --- | --- | --- | --- |
| 253 | VA | 9300 | 28 | 1970 | 9272 | 1942 |
| 254 | VA | 486 | 1 | 1970 | 485 | 1969 |
| 255 | VA | 1070 | 1 | 1970 | 1069 | 1969 |
| 256 | VA | 1001 | 1 | 1220 | 1000 | 1219 |
| 257 | VA | 1135 | 29 | 1970 | 1106 | 1941 |
| 258 | VA | 1201 | 1 | 1970 | 1200 | 1969 |
| 259 | VA | 11250 | 1 | 1970 | 11249 | 1969 |
| 260 | VA | 1821 | 51 | 1970 | 1770 | 1919 |
| 261 | VA | 16251 | 1 | 1970 | 16250 | 1969 |
| 262 | VA | 62500 | 34 | 1970 | 62466 | 1936 |
| 263 | VA | 736 | 1 | 1970 | 735 | 1969 |
| 264 | VA | 12000 | 6 | 1970 | 11994 | 1964 |
| 265 | VA | 750 | 7 | 1970 | 743 | 1963 |
| 266 | VA | 1876 | 1 | 1970 | 1875 | 1969 |
| 267 | VA | 511 | 84 | 1905 | 427 | 1821 |
| 268 | VA | 1600 | 3 | 1970 | 1597 | 1967 |
| 269 | VA | 1285 | 40 | 1970 | 1245 | 1930 |
| 270 | VA | 1274 | 174 | 1970 | 1100 | 1796 |
| 271 | VA | 499 | 3 | 1970 | 496 | 1967 |
| 272 | VA | 12000 | 4 | 1970 | 11996 | 1966 |
| 273 | VA | 67749 | 1 | 1085 | 67748 | 1084 |
| 282 | VA | 1000 | 3 | 1885 | 997 | 1882 |
| 283 | VA | 414 | 224 | 1885 | 190 | 1661 |
| 284 | VA | 32250 | 1 | 1885 | 32249 | 1884 |
| 285 | VA | 11400 | 6 | 1885 | 11394 | 1879 |
| 286 | VA | 30747 | 2 | 1885 | 30745 | 1883 |
| 287 | VA | 9651 | 1 | 1885 | 9650 | 1884 |
| 288 | VA | 1135 | 1 | 1885 | 1134 | 1884 |
| 289 | VA | 3170 | 2 | 1885 | 3168 | 1883 |
| 290 | VA | 236 | 172 | 1885 | 64 | 1713 |
| 253 | VAM | 11391 | 2 | 2060 | 11389 | 2058 |
| 254 | VAM | 17400 | 1 | 2060 | 17399 | 2059 |
| 255 | VAM | 4366 | 1 | 2060 | 4365 | 2059 |
| 256 | VAM | 2691 | 1 | 1310 | 2690 | 1309 |
| 257 | VAM | 2611 | 1 | 2060 | 2610 | 2059 |
| 258 | VAM | 6101 | 1 | 1855 | 6100 | 1854 |
| 259 | VAM | 2351 | 1 | 2060 | 2350 | 2059 |
| 260 | VAA | 400 | 1 | 2060 | 399 | 2059 |
| 261 | VAM | 7076 | 1 | 2060 | 7075 | 2059 |
| 262 | VAM | 15851 | 1 | 2060 | 15850 | 2059 |
| 264 | VAA | 5801 | 1 | 2060 | 5800 | 2059 |
| 265 | VAM | 2115 | 68 | 2060 | 2047 | 1992 |
| 266 | VAM | 2396 | 1 | 2060 | 2395 | 2059 |
| 267 | VAM | 2895 | 11 | 2060 | 2884 | 2049 |
| 268 | VAM | 354 | 1 | 2060 | 353 | 2059 |
| 269 | VAM | 1325 | 3 | 2060 | 1322 | 2057 |
| 270 | VAM | 5450 | 805 | 2060 | 4645 | 1255 |
| 271 | VAM | 4336 | 1 | 2060 | 4335 | 2059 |
| 272 | VAM | 13400 | 31 | 2060 | 13369 | 2029 |
| 273 | VAM | 17800 | 1 | 2060 | 17799 | 2059 |
| 282 | VAM | 1376 | 1 | 2060 | 1375 | 2059 |
| 283 | VAM | 6851 | 69 | 2060 | 6782 | 1991 |
| 284 | VAM | 16251 | 1 | 2060 | 16250 | 2059 |
| 285 | VAM | 3825 | 1 | 2060 | 3824 | 2059 |
| 286 | VAM | 3701 | 1 | 2060 | 3700 | 2059 |
| 287 | VAM | 4501 | 1 | 2060 | 4500 | 2059 |
| 288 | VAM | 5701 | 1 | 1890 | 5700 | 1889 |
| 289 | VAM | 18901 | 1 | 1890 | 18900 | 1889 |
| 290 | VAM | 10550 | 245 | 1885 | 10305 | 1640 |
